# A multimodal dataset for precision oncology in head and neck cancer

**DOI:** 10.1101/2024.05.29.24308141

**Authors:** Marion Dörrich, Matthias Balk, Tatjana Heusinger, Sandra Beyer, Hassan Kanso, Christian Matek, Arndt Hartmann, Heinrich Iro, Markus Eckstein, Antoniu-Oreste Gostian, Andreas M. Kist

## Abstract

Head and neck cancer is a common disease and is associated with a poor prognosis. A promising approach to improving patient outcomes is personalized treatment, which uses information from a variety of modalities. However, only little progress has been made due to the lack of large public datasets. We present a multimodal dataset, HANCOCK, that comprises monocentric, real-world data of 763 head and neck cancer patients. Our dataset contains demographical, pathological, and blood data as well as surgery reports and histologic images. We show its potential clinical impact in a multimodal machine-learning setting by proposing adjuvant treatment for previously unidentified risk patients. We found that especially the multimodal model outperformed single-modality models (area under the curve (AUC): 0.85). We believe that HANCOCK will not only open new insights into head and neck cancer pathology but also serve as a major source for researching multimodal machine-learning methodologies in precision oncology.

## Introduction

Head and neck cancer is the seventh most common malignancy worldwide [1]. Patients diagnosed with head and neck cancer have a poor prognosis [2]. Despite recent advances in diagnostics and treatments, such as immunotherapy, the 5-year survival ranges only between 25% and 60% [3]. The most common head and neck cancer develops in several locations, e.g. the oral cavity, pharynx, or larynx, and is derived from squamous cells, i.e. originates from the mucosal epithelium lining the inner areas of these sites. The cancer often spreads to regional lymph nodes, which further worsens the prognosis of affected patients [4].

After assessing the medical history and physical examination, a panendoscopy with biopsy is usually performed to confirm the diagnosis. The pathological analysis of tissue samples is crucial for determining the histological entity. Additionally, lymph nodes are examined for possible metastases. Surgery is one of the most important pillars of treatment for head and neck cancer. Local surgery is often sufficient for lower-stage cancer, while adjuvant treatment such as radiotherapy or radiochemotherapy is required for higher stages [5]. Despite many advances in diagnostics, the treatment choice still depends mainly on the stage of the disease that is mainly determined by the size of the tumor [5, 6]. However, research showed that cancer is highly diverse among patients [7] and therefore requires precision oncology. The key to this personalized treatment is the establishment of reliable and predictive biomarkers. Initiatives such as The Cancer Genome Atlas (TCGA) have already achieved a better understanding of the genetic and molecular characteristics of many types of cancer [8].

However, very few biomarkers are currently used in routine head and neck cancer treatment. A positive prognostic biomarker is the association with human papillo-mavirus (HPV) in oropharyngeal carcinomas [9]. Ongoing research aims to explore if their treatment can be de-escalated to reduce toxicity [10]. Furthermore, the expression of programmed death ligand 1 (PD-L1) can be assessed to identify patients who may benefit from immune checkpoint inhibitors such as pembrolizumab, and remains the only applied predictive biomarker for now [11]. However, more reliable biomarkers need to be established to enable a truly personalized treatment. Although information from a large variety of sources is routinely acquired, its full potential cannot be realized for data-driven exploration yet. Careful data curation and multimodal integration are required to unravel complex data dependencies. We hypothesize that a lack of such large, multimodal, publicly available datasets hinders the research of predictive biomarkers for head and neck oncology.

To our knowledge, existing head and neck cancer datasets only have a limited number of cases or have inconsistent metadata [12–15]. For example, a study focusing on radiomics included data from 288 cases while only selecting oropharyngeal carcinomas [15]. Another dataset focusing on proteomics includes radiology and histopathology data but is limited to 122 cases [13]. Comprehensive data including clinical, genomic, and histopathologic data has been collected on TCGA from more than 500 cases to date, however, the multicenter data is very heterogeneous [12, 16]. To address these issues, we collected monocentric, retrospective data from more than 700 head and neck cancer patients. We built a comprehensive dataset from multimodal data including demographics, blood data, surgery reports, pathologic data, and histologic images. These include Whole Slide Images (WSIs) with routine hematoxylin and eosin (HE) staining and Tissue Microarrays (TMAs) with staining for several immune cell populations. In this work, we aimed to explore and provide reproducible strategies for multimodal integration and analysis. We aimed to predict patient outcomes and investigate adjuvant treatment choices using multimodal Machine Learning (ML) strategies to show the impact of multimodal data integration for head and neck oncology.

## Results

### Compilation of a multimodal dataset from a head and neck cancer cohort

Patient diagnoses and treatment decisions are rarely based on a single modality; hence, artificial intelligence (AI) models intended to assist clinicians should adopt a holistic approach, incorporating multiple data sources. Training such models requires extensive and diverse patient data, which is often scarce. To address this, we have aggregated a comprehensive dataset, HANCOCK (Head And Neck Cancer dataset), which consists of real-world data from 763 patients. In detail, we collected, cleaned, and harmonized routinely acquired monocentric data from patients diagnosed with oral cavity, oropharyngeal, hypopharyngeal, and laryngeal cancer. We integrated different modalities including demographics, blood data, pathology reports, surgery reports, and histologic images, as shown in Figure 1A. We provide an overview and easy, public access to the individual patient data for convenient manual exploring at www.hancock.research.fau.eu with support of the FAUDataCloud.

**Fig. 1.**
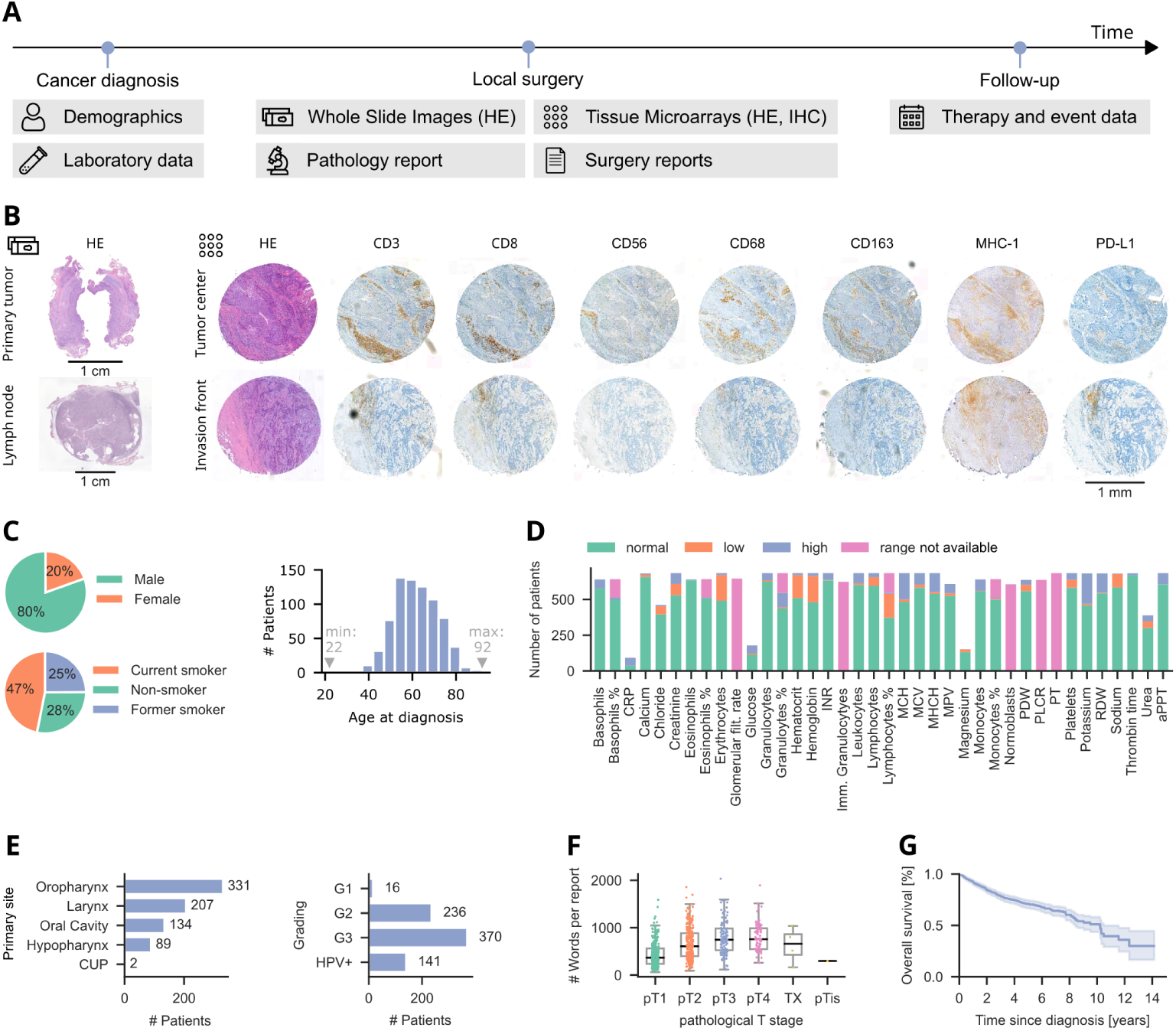
Overview of the multimodal head and neck cancer dataset. (A) Data sources. For cancer diagnosis, demographics were assessed and blood tests were performed. In the ablative surgery, tissue samples were obtained and the pathological report was written. The dataset also features information about the treatment choice, events, and survival. (B) Image data of a patient. Shown are Whole Slide Images of the primary tumor and lymph node with hematoxylin and eosin (HE) staining and Tissue Microarray cores from the tumor center and invasion front with HE and immunohistochemistry (IHC) staining. (C) Demographical data, shown as the number of patients per sex, smoking status, and age at initial diagnosis. (D) Laboratory data. Shown is the number of patients for which each parameter is available. The colors indicate values inside or outside of the normal range. (E) Primary tumor site or CUP (cancer of unknown primary) and grading from the pathology report. HPV-associated carcinoma was not graded. (F) Number of words in each German surgery report grouped by pathological T stage. (G) Kaplan-Meier plot of overall survival with 95% confidence interval shown as shaded error.

A core strength of HANCOCK is its rich base of imaging data: HE-stained WSIs of the primary tumor are available for 701 out of 763 patients. We provide also manual annotations of tumor regions in these WSIs, as shown in Supplementary Fig. S1. In addition, 396 HE-stained slides of adjacent lymph nodes were included. Each patient contains at most 32 TMAs, which reflect two cores, eight stains, and two locations. Each core is stained with either HE or immunohistochemistry (IHC) markers, such as CD3 and PD-L1. Figure 1B shows exemplarily the available imaging data for a single patient. For each patient, the pathology report was included in a structured format. These cover tumor characteristics such as the primary site or grading (see Figure 1E) crucial for selecting a suitable treatment. Additional characteristics such as tumor staging, resection margin, and infiltration depth are summarized in Supplementary Fig. S2.

As shown in Figure 1C, 80% of the patients in the dataset are males and 72% are former or current smokers. The median age is 61 years. Thus, our patient cohort reflects the current demographics of head and neck cancer [1], which is beneficial for generalizing our findings to a broader population. The laboratory data includes the complete blood count as well as coagulation parameters, electrolytes, renal function parameters, and C-reactive protein. Figure 1D shows for how many patients the individual parameters are available and how many of the measured blood parameters are in the normal or abnormal range.

The incorporation of treatment information and temporal event data allows an in-depth analysis of the underlying relationships. To this end, we extracted and de-identified plain text descriptions of the surgery and medical history from text documents. Figure 1F illustrates the length of surgery reports, which seems to increase with the pathological T stage. All German text files were translated into English to improve their accessibility (see Methods). OPS codes (German procedure classification) define the medical procedures applied. We also extracted ICD codes (International Statistical Classification of Diseases and Related Health Problems) of the German version ICD-10-GM from the text documents. The ICD codes allow a detailed classification of malignancies and their sites. The most frequent ICD codes were C10.8 and C32.0, as shown in Supplementary Fig. S3D. C10.8 corresponds to a malignant neoplasm in overlapping regions of the oropharynx and C32.0 corresponds to a malignant neoplasm of the glottis [17]. We believe that ICD coded will allow easy subsampling of the full dataset.

In HANCOCK, each patient is tracked from the time of initial diagnosis to either the end of follow-up or death, with follow-up periods lasting as long as 14 years (see Supplementary Fig. S4). This enables the examination of temporal information, for example in the form of treatment timelines (see Supplementary Fig. S5) and survival analyses. Figure 1G shows the overall survival of all patients in the HANCOCK dataset. Survival curves with additional information such as the number of censored patients can be found in Supplementary Fig. S6D and survival curves grouped by primary site, stage, and grading are shown in Supplementary Fig. S6A-C. The 5-year survival rate in our cohort is 77.3%.

Overall, the dataset features a great variety of modalities for a large patient cohort (763 cases), which resembles the global demographics of head and neck cancer.

### Multimodal data integration allows prediction of clinical outcomes

After carefully aggregating the patient data, we were next interested in investigating the overall patient collective. To better understand the complex patient data, we encoded information from each modality individually and concatenated these encodings into vectors, termed multimodal patient vectors, as shown in Figure 2A. Given the high-dimensional nature of these vectors (the multimodal patient vectors contain 103 dimensions each, see Methods), these patient-centered features can hardly be examined or interpreted by humans. Therefore, we applied Uniform Manifold Approximation and Projection (UMAP) to these vectors to project them into a lower, two-dimensional space, as shown in Figure 2B and C. In Supplementary Fig. 7, we provide a comprehensive overview of incorporated features and their distribution in the UMAP projection.

**Fig. 2.**
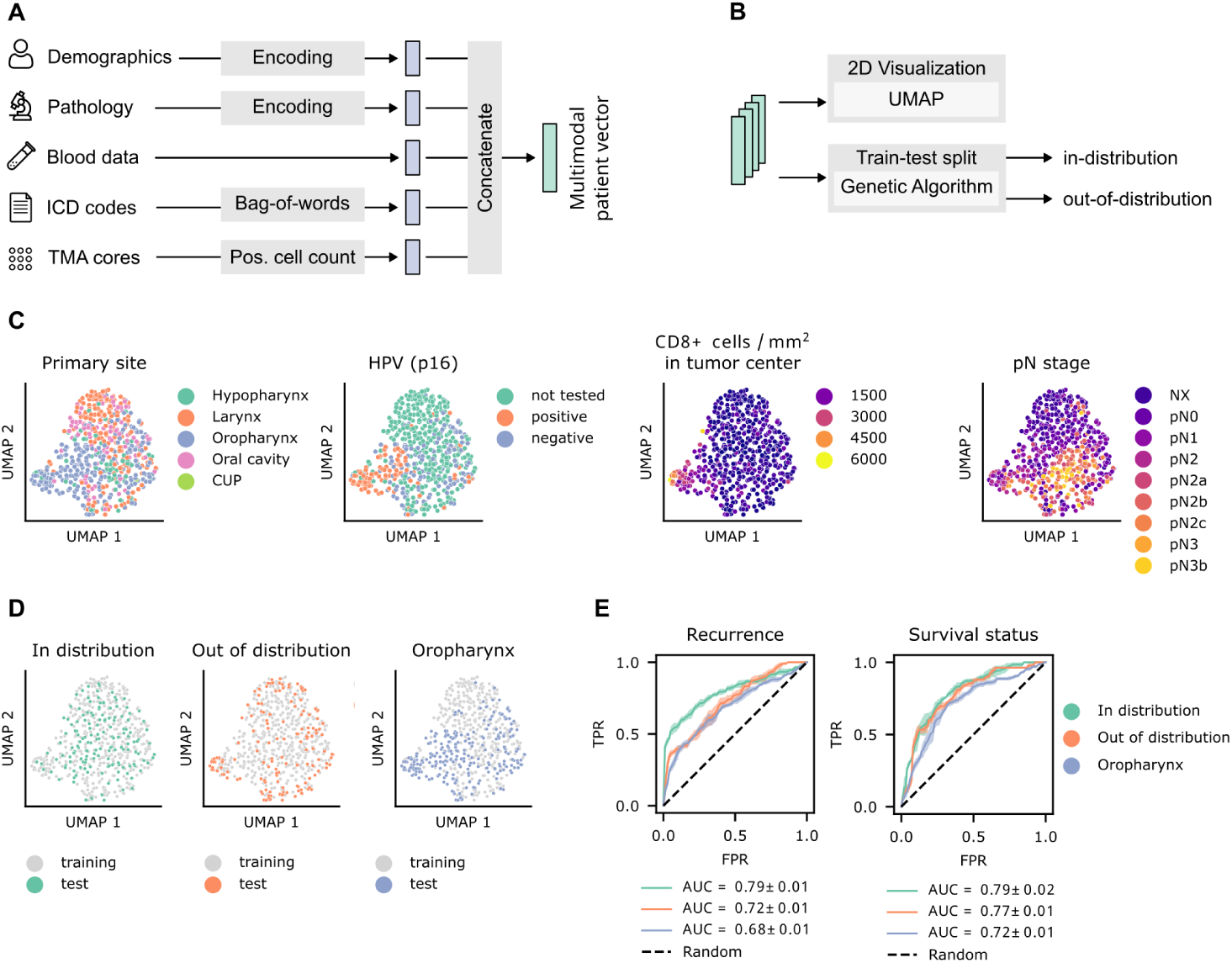
Multimodal embeddings. (A) For each patient, information from distinct modalities were encoded and concatenated to multimodal patient vectors. (B) We applied Uniform Manifold Approximation and Projection (UMAP) to visualize the vectors in 2D and we implemented a genetic algorithm to create two test datasets, one in the distribution of the training data and one out of the distribution. (C) Visualization of two-dimensional embeddings, colored by features of the encoded data. (D) UMAP plots of three different train-test splits (E) Receiver-operating characteristics (ROC) curves of a Random Forest classifier for the three splits and two prediction tasks. The mean values and standard deviations of the ROC curves and Area under the Curve (AUC) scores are shown. The colors correspond to the different splits in D.

Subsequently, we sought to identify distinct patient clusters using these multimodal patient vectors. We hypothesized that similar patient groups would converge within specific areas of the two-dimensional UMAP projection. Our findings confirm this hypothesis, as we observed that patients sharing particular characteristics tended to form distinct clusters. For instance, patients diagnosed with HPV-positive oropharyngeal carcinoma often exhibited a high density of CD8+ cells, as illustrated in Figure 2C. Additionally, our analysis revealed that both CD3+ and CD8+ cell densities at the tumor center and the invasion front were notably higher in patients who did not experience recurrence compared to those who did (Supplementary Fig. S8). These observations are consistent with prior studies in head and neck oncology [9], underscoring the relevance and accuracy of the HANCOCK dataset.

We aimed to investigate whether ML models could predict clinical outcomes, i.e. recurrence and survival status, using the encoded multimodal data. We were also interested in defining different hold-out test datasets that would allow a robust estimation of a model’s performance. To this end, we defined three data splits that divide the cases into one training and one test set. We hypothesize that the performance of models can be over- or under-estimated depending on how similar the test data is to the training data, especially in a complex, high-dimensional, and multimodal setting as in our case. To address and investigate this issue, we implemented a genetic algorithm to automatically define two dataset splits based on multidimensional features. The algorithm uses evolutionary optimization to find (i) cases that follow the overall distribution (”in distribution”) or (ii) cases that lie outside the distribution and are maximally dissimilar to each other (”out of distribution”). In both settings, the genetic algorithm preserves the distribution of target classes (recurrence and survival status) in the resulting training and test sets, which is important for model evaluation [18]. The respective class distributions are shown in Supplementary Fig. S9C-D. Additionally, we defined a third split where all patients with a carcinoma located in the oropharynx were assigned to the test dataset, rendering it very dissimilar and biased to the training data. These three training/test data splits are highlighted in the UMAP representation in Figure 2D.

Next, we trained an ML model, namely a Random Forest classifier, to predict the recurrence and survival status of each patient by using the multimodal patient vectors as inputs. This corresponds to an early fusion approach since the modality vectors are first concatenated and then used to train a single model [19]. Figure 2E shows the performance of the classifiers for the previously mentioned train-test splits (see Figure 2D for reference). As expected, the model had difficulty predicting patient outcomes for the test dataset consisting of cases with oropharyngeal carcinoma, a primary site that the model has not seen during training. This is highlighted by the lowest Area Under the Curve (AUC) score as shown in Figure 2E compared to the other test sets. In accordance with our hypothesis, the classification performance was higher for the ”in distribution” than the ”out of distribution” test dataset as shown in Figure 2E. Overall, we can provide evidence that multimodal ML models follow expected ML behavior and were able to successfully estimate the prognosis of patients, achieving a maximum average AUC score of 0.79 for both recurrence and survival prediction.

### Multimodal machine learning enables improved adjuvant treatment selection

An important choice in oncologic therapy is whether an adjuvant treatment is required for a given patient. That means, identifying risk patients that benefit from an adjuvant therapy is crucial. We analyzed the HANCOCK patient cohort and found that some patients did not receive adjuvant treatment, but eventually had a recurrence or deceased (Figure 3A), suggesting that exactly this patient collective are risk patients who would have potentially benefited from adjuvant therapy. We assume that all other patients in our dataset received appropriate treatment to the best of the treating physicians’ knowledge. We then were interested in how the potentially unidentified risk patients would have been classified (adjuvant therapy needed yes/no) by a multimodal ML model. Hence, we assigned these cases to a hold-out test dataset (Figure 3A). The remaining cases, i.e. cases with adjuvant therapy and cases without adjuvant therapy and no recurrence or death, were assigned to a training dataset.

**Fig. 3.**
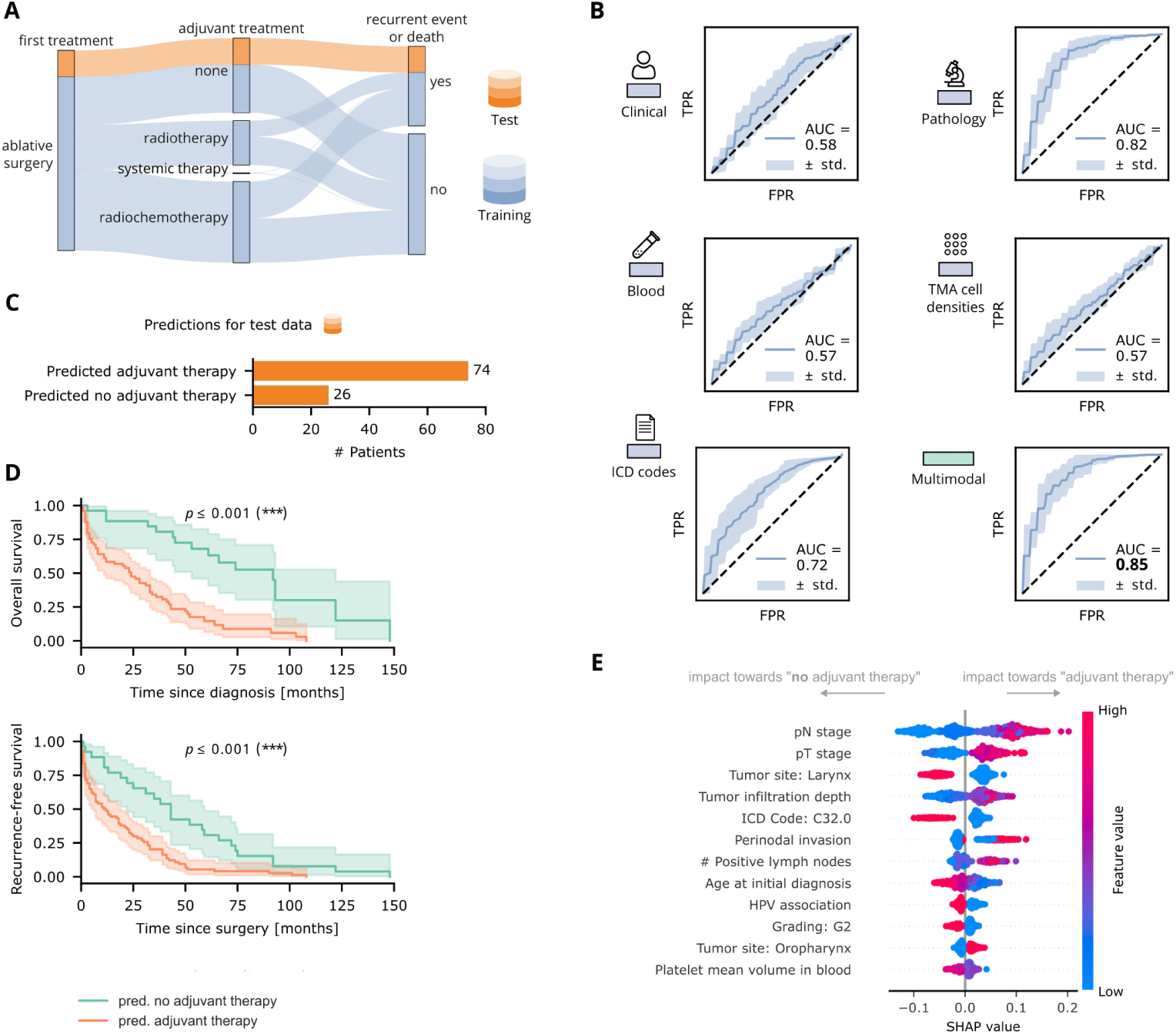
Prediction of treatment choice. (A) Patients who did not receive adjuvant therapy but did have a recurrence or deceased within 5 years (highlighted in orange) were assigned to the test dataset. All other patients were assigned to the training dataset. (B) Receiver-operating characteristic (ROC) curves of Random Forest classifiers trained on single-modal and multimodal data using 10-fold cross-validation with the mean Area Under the Curve (AUC). (C) The multimodal model predicted adjuvant therapy for 74% of cases in the test dataset. (D) Kaplan-Meier curves for the test dataset, with patients grouped by predictions. The log-rank test was used. (E) SHAP summary plot for model interpretability, showing the 12 most important features of the multimodal model (trained on the full training data), computed for all validation folds.

First, we evaluated the benefits of multimodality vs. single modalities. Therefore, we trained ML models on the multimodal patient vectors and each of the modalities separately. These modalities include clinical, pathological, and blood data as well as the density of CD3- and CD8-positive cells and ICD codes. Figure 3B shows the corresponding average Receiver-operating characteristic (ROC) curves using 10-fold cross-validation. As shown in Figure 3B, the classifier integrating the multimodal data outperformed all single-modality classifiers with a mean AUC score of 0.85. This finding is in line with previous works that have shown the superior performance of ML models trained on several modalities compared to data with limited information from a single source [20, 21]. Out of the single-modality models, the classifier trained on pathological data achieved the highest mean AUC score of 0.81, as shown in Figure 3B.

We next trained a classifier on the full, multimodal training data. Figure 3C shows the predictions of this trained model for the hold-out test dataset, i.e. the potential risk patients (orange cohort in Figure 3A). Figure 3C reveals that the multimodal ML classifier suggested an adjuvant therapy for 74 out of 100 cases. Furthermore, Figure 3D shows that the 74 patients for whom an adjuvant treatment was proposed, were high-risk patients i.e. their probability of recurrence-free survival and overall survival were significantly lower than for the other 26 patients (p *≤* 0.001, log-rank test).

The incorporation of ML in clinical practice is often hindered by a lack of explainability and its ”black box” nature [22]. To improve the interpretability of our multimodal approach, we obtained SHAP values that explain the impact of individual features on the model output [23]. Figure 3E shows the twelve most important characteristics in a summary plot. The four features with the highest impact were pathological features, agreeing with the high AUC score of the pathological model in Figure 3B. For example, a high pathological N and T stage led to a higher probability of predicting an adjuvant treatment. These two features had the greatest impact on the predictions, which is consistent with the fact that the treatment choice mainly depends on the stage of the disease [5]. However, adjuvant treatment was not likely to be predicted for laryngeal carcinomas and glottic carcinomas in particular, as shown in Figure 3E as the ICD code C32.0 stands for malignant neoplasms of the glottis [17]. A demographic feature, namely the age at initial diagnosis, also had a high impact on the outputs; With increasing age, the need for an adjuvant therapy became less likely. Furthermore, an HPV association had a high impact on not predicting adjuvant treatment, which is consistent with results showing that HPV-positive patients have a better prognosis [9] (see survival grouped by HPV status in Supplementary Fig. 6D). Taken together, our results suggest that multimodal models can integrate more valuable information than single modality models, and could be useful for assisting in adjuvant treatment selection: in this case, 3 out of 4 patients would have potentially benefited from an adjuvant therapy questioning current clinical guidelines and suggesting the incorporation of multimodal ML models. We showed that our ML model relied on the stage and also on a variety of other characteristics such as infiltration depth, perinodal invasion, age, and HPV association.

### Treatment choice prediction using immunohistochemistry images

Computer vision approaches on histopathological image data have shown promising results in a variety of oncology settings [21, 22, 24]. We were interested if the image data in the HANCOCK dataset is as well suited for multimodal data integration. Using the dataset split shown in Figure 3A, we explored an approach for integrating image features and the encoded tabular data to train a convolutional, deep neural network. To this end, we analyzed the TMAs taken from the tumor center. Each TMA contains multiple samples and two cores were available for each patient, as shown in Figure 4A. We extracted a single 1024*×*1024 µm tile from each TMA core. Figure 4B shows that we used TMAs stained with seven distinct IHC markers and the standard HE stain. All tiles were fed to a VGG16 pre-trained on ImageNet to extract high-level features [25, 26]. The features were ”deep texture representations” of the images, following the technique of Komura et al. [27]. We found that there was a relationship between these image representations and the computed cell density of CD3- and CD8-positive cells, as shown in Supplementary Fig. S10.

**Fig. 4.**
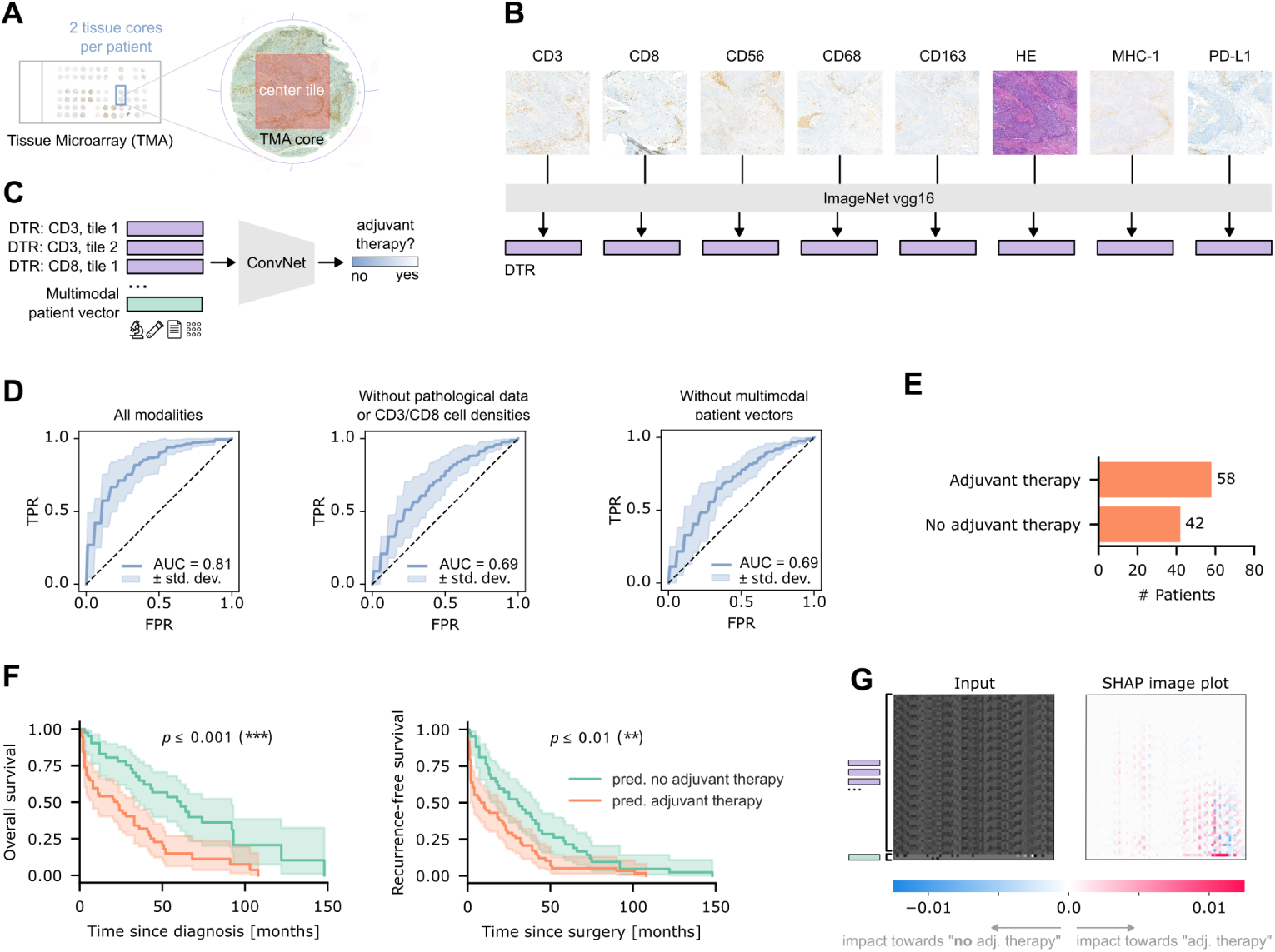
Combining multimodal feature vectors with image embeddings. (A) Tile extraction. From each TMA cores (two cores per patient), a tile was extracted from the center. (B) Vgg16, pre-trained on ImageNet, was used to extract deep texture representations [27] from tiles extracted from TMAs with 8 distinct markers, of which 7 are immunohistochemistry markers. (C) Stacking of features to train a Convolutional Neural Network for treatment choice prediction. (D) Receiver-operating characteristic (ROC) curves from 10-fold cross-validation. In the left plot, all modalities were used. In the center plot, pathological features and cell densities were excluded. In the right plot, the multimodal patient vectors were excluded. (E) Predictions for the test dataset. (F) Kaplan-Meier curves for the test dataset, with patients grouped by predictions (G) Visual explanation using SHAP values for a test sample.

For each patient, a two-dimensional embedding was created by stacking the image features and the multimodal patient vectors (see Methods). The resulting image-like embeddings were used to train a Convolutional Neural Network (CNN, see Figure 4) to the same task as in Figure 3.

Figure 4D shows that the network achieved a mean AUC of 0.81 in 10-fold cross-validation. Thus, it did not outperform the ML model trained on the high-level multimodal patient vectors alone (compare Figure 3C) but performed in a similar range. We hypothesized that some parts of the multimodal feature vectors contained information that overlapped with the information in the image embeddings, namely the structured pathological data and cell densities, which are derived from the histopathological imaging data. Figure 4D shows that models lacking these features resulted in a decrease in the classification performance. We found that the CNN was still able to reach a mean AUC of 0.69 on the image representations alone which indicates that valuable information was contained in the extracted features.

The network predicted an adjuvant treatment for 58 out of 100 cases, see Figure 4E. The respective Kaplan-Meier curves (see Figure 4F) show that the overall and recurrence-free survival probability was significantly lower in patients, for whom the model suggested adjuvant therapy (p *≤* 0.001 and p *≤* 0.01, log-rank test).

Next, we were interested in analyzing the impact of different modalities on the CNN’s predictions. We generated SHAP image plots to create visual explanations. An example is shown in Figure 4G. We found that patterns within the image embeddings as well as individual features in the multimodal patient vectors were highlighted. This indicates that stacking extracted image features and encoded tabular features might be a valuable approach to multimodal Deep Learning with the advantage of being computationally inexpensive. Overall, we were able to integrate image data using a simple early fusion approach to train a deep neural network, yielding promising results.

## Discussion

In this study, we provide a novel monocentric dataset - HANCOCK - comprising 763 patients with multimodal data. The modalities include demographical, pathological, and blood data, WSIs from primary cancer and lymph nodes, and TMAs with IHC staining. We show that the dataset is rich and diverse, and not biased towards a single domain (Figures 1 and 2). By integrating multimodal data through diverse machine and deep learning approaches, we can show that this allows better prediction of survival and recurrence (Figure 2F), as well as providing a superior choice for adjuvant therapy across AI technologies (Figures 3 and 4). With our transparent and open approach, we hope the HANCOCK dataset will fuel further developments in multimodal data integration and head and neck oncology. By reproducing previous findings, such as the predictive behavior of HPV and PD-L1, we believe that HANCOCK will be very useful in biomarker discovery and validation.

We found some limitations in our work, which can be addressed in future studies. For example, we did not integrate WSIs of the primary tumors or lymph nodes to train deep neural networks. Instead, we focused on TMA tiles as inputs since they provide information about distinct immune cell populations and are also available with routine HE staining. Another advantage of using the TMAs was that they were taken specifically from the tumor region. However, integrating the HE-stained WSIs additionally could further improve the prediction of clinical outcomes or treatment choices since multiple studies have shown that neural networks trained on WSIs alone can predict risk or prognosis, for example using Multiple-Instance-Learning [28–30].

We further relied consistently on an early fusion approach for training any multimodal AI in our study. This means first fusing the features of distinct modalities and then training a single model, which is recommended as an initial strategy [19]. Future studies should also evaluate if the classification performance could be improved using joint fusion, where neural networks are not only used as feature extractors but are also trained in the process [19]. An in-depth comparison of different methods for extracting and fusing features, especially from our comprehensive histologic image data, could be very beneficial.

We extracted ICD codes from the surgery reports and integrated them into multimodal embeddings using bag-of-words. However, we did not incorporate the plain texts themselves. Since the surgery reports describe the tumor resection in detail and could potentially provide additional information about the severity of the disease, they could be further explored. For example, text embeddings could be extracted using a pre-trained transformer and integrated into the multimodal vectors [20].

Our Machine Learning models are limited to binary classification, however, other options could be explored using the available event data. For example, a regression model could be implemented to predict the time to events such as recurrence or death. Moreover, models could be trained to predict risk scores using a loss function such as Cox partial likelihood loss as proposed by Chen et al. [21].

In this work, the densities of CD3- and CD8-positive cells were computed from the TMAs. We analyzed these regarding their relationship to clinical outcomes (see Supplementary Fig. S8) and integrated them in the multimodal vectors for ML model training (see Figure 2A). In the future, immune cells expressing the markers CD56, CD163, PD-L1, and MHC-1 as available in HANCOCK could be analyzed as well and integrated for ML model training accordingly.

A limitation of our multimodal ML model for treatment prediction is that it could not account for all possible reasons for deciding against adjuvant therapy. For example, no data about patient refusal or comorbidities was available. Hence, collecting more detailed information about the process of treatment selection could be beneficial.

It has been shown that tissue or cell detection and subsequent classification can enable the investigation of quantitative biomarkers [31, 32]. Therefore, annotations of the histologic images in our dataset could be beneficial for biomarker discovery. We already provide manual annotations of tumor regions in the WSIs of the primary tumor. However, these annotations were done sparsely instead of exhaustively and they were not done by pathologists. We aim to extend HANCOCK in the future, for example by creating high-quality annotations of distinct cell types. To this end, we could leverage Deep Learning models and existing manual annotations of nuclei. The annotation or segmentation of larger tissue regions could also be considered and incorporated into the dataset. Further, combining molecular data with histopathological data is a promising approach [33]. Hence, we aim to further integrate genomic or transcriptomic data, to increase the long-term impact of the dataset.

Finally, HANCOCK allows the possibility to explore the concept of digital twins, a digital representation of cancer patients, that could improve decisions in cancer care [34]. We implicitly used this concept in the training/test data split (Figure 2) to compute the cosine similarity between patients to ensure a specific distribution of patients in a given subset (see Methods).

## Methods

### Data collection

The data was acquired from the Department of Otorhinolaryngology and Head and Neck Surgery and from the Pathological Institute of the University Hospital in Erlangen. All data was collected and published following the local ethics committee vote (#23-22-Br). Retrospective, multimodal data was gathered from patients who were diagnosed with head and neck cancer between 2005 and 2019. Only patients who had a curative first treatment were included. The modalities in our dataset can be categorized into image data (histopathological images), structured data (clinical, pathological, and blood data), and free text (surgery reports). Supplementary Fig. S11 shows the available and missing data types for all patients.

Tissue samples of the respective patients were collected from the pathological archive of the University Hospital in Erlangen. The samples originate from the primary tumor and, if present, positive lymph nodes that had been resected. The tissue samples had been fixed in formalin, embedded in paraffin, and routinely stained with HE. The 709 primary tumor sections were scanned using a 3DHistech P1000 at 82.44*×* magnification. A single slide was available for 701 cases whereas two slides were available for eight cases. The 396 lymph node sections were scanned using an Aperio Leica Biosystems GT450 at 40*×* magnification and using 3DHistech P1000 at 51.42*×* magnification. All digitized WSIs were stored in the pyramidical Aperio file format (.svs). Additionally, TMAs were created from the paraffin-embedded primary tumor blocks. The TMA cores with a diameter of 1.5 mm were extracted from the tumor center and the tumor invasion front. They were stained using HE and they were stained for specific immune cell populations using the IHC markers CD3, CD8, CD56, CD68, CD163, PD-L1, and MHC-1. CD3-positive cells represent T cells, CD8-positive cells represent cytotoxic T cells, and CD56-positive cells represent natural killer cells. CD68 and CD163 were used to detect monocytes and macrophages. PD-L1 plays a major role in regulating the immune response. It is expressed by tumor cells to deactivate cytotoxic T cells and is a target for immunotherapy [35]. The major histocompatibility complex class I (MHC-1) displays antigens to cytotoxic T cells and is also important for determining the prognosis and treatments involving immunotherapy [36]. From each patient, at least two cores were collected per origin and marker. This resulted in 368 TMAs, each with cores arranged in 12 rows by 6 columns. The TMAs were scanned using a 3DHistech P1000 at 82.44*×* magnification.

Structured pathological data originating from the analysis of the primary tumor and lymph node sections was harmonized and compiled in tabular format. It includes comprehensive information such as the cancer site, staging, grading, and histologic type. The clinical data includes each patient’s age, sex, and smoking status. It further contains information and timestamps of events such as treatments, recurrence, progress, metastasis, or death. The data was collected from the hospital information system and by screening various documents such as general and radiotherapy records. Blood test results of the corresponding patients in a range of 14 days around local surgery were retrieved from the hospital’s archive. Each measurement was accompanied by the parameter’s name, group, unit, and LOINC code (Logical Observation Identifiers Names and Codes) [37].

Surgery reports were collected by filtering the hospital’s database by patient identifiers and time range. Reports of patients diagnosed in 2006 were not available, as reports were not entered into the database until 2007. The surgery reports follow a template that includes the medical history and report in the document’s body and metadata in the header. All documents were compiled into a .pdf file.

### Data preprocessing

The data was anonymized by assigning a unique, consecutive ID (”001” to ”763”) randomly to each patient. Our data is patient-centered. This means that each WSI, each core in a TMA, each surgery report, and each entry in the structured data is mapped to a single patient ID. The preprocessing steps for each data modality are described in the following.

TMAs and WSIs were converted from the manufacturer’s file format (.mrxs) to the pyramidical Aperio file format (.svs). An Aperio SVS file contains a macro image and a label image. The label image in particular contains potentially identifying information. Therefore, we anonymized the files by removing the label images, i.e., by replacing the image with zeros. To allow the mapping of each TMA core to the corresponding patient, we created TMA maps in .csv format (comma-separated values) that can be imported into QuPath.

We identified the most important clinical and pathological features and ensured that these were complete for all patients. We performed data cleaning to remove inconsistent or redundant data. For patients with more than one entry in the clinical table, we kept the entry with the earlier diagnosis date. Each following entry was discarded because it reported a recurrence of the disease rather than the initial diagnosis. We de-identified the clinical and pathological data by removing all names and dates. The year of the initial diagnosis was retained, but its date was removed. For anonymization purposes, all dates of events were replaced by the number of days since the initial diagnosis. This way, the timeline from the diagnosis to the end of treatment could still be reconstructed. We corrected spelling errors, summarized and harmonized table entries, and assigned self-explanatory labels. The tables were finally converted into Javascript-Object Notation (JSON). Descriptions of all fields in the JSON files with their data types and possible values were summarized in data dictionaries, shown in Supplementary Tables S1, S2, and S3.

The results of blood tests were available as structured, tabular data. We first filtered the data to select values that were measured at specified units, excluding intensive care units. For each patient, we chose a single pre-operative measurement of each parameter. To this end, we selected the latest available measurement before the surgery date because relevant blood tests are usually performed one to three days before. If no pre-operative value was available, the value from the surgery day itself was selected. The number of available measurements for these time points is shown in Supplementary Fig. S12. The complete blood count, coagulation parameters, electrolytes, and renal function parameters were routinely assessed. Additional parameters were calcium, magnesium, glomerular filtration rate, and glucose. Although it was only available for 94 patients, we included C-reactive protein (CRP) since elevated CRP levels are associated with poor prognosis in patients with head and neck cancer [38]. The blood dataset was converted to JSON format.

The surgery reports were first converted from .pdf to .txt format. Each document had a header containing the operating clinicians, treatment date, the patient’s name, and identifiers such as the admission number. The header additionally contained OPS codes and ICD codes. We used regular expressions in Python to search for keywords and obtain relevant data. This way, we extracted ICD codes, OPS codes, and the medical history along with the surgery report itself. We selected reports from the first treatment date, i.e. from the local surgery, and discarded all others. Most patient names had already been masked when they had been entered into the system. However, many texts contained names of operating clinicians. Therefore, we used regular expressions to substitute any names following medical or academic titles. Additionally, we performed a search using regular expressions and lists of all names of patients and clinicians. Finally, the reports and medical histories were screened manually for any remaining identifying information. Patient names, clinician names, locations, and dates were replaced by placeholders. The number of replaced terms is shown in Supplementary Table 4. The documents were saved to plain text (.txt) files. Additionally, we translated all surgery reports, and medical histories from German to English using the DeepL API [39]. For translating short descriptions to English, we used ChatGPT (GPT-3.5) [40]. For convenience, HANCOCK contains the German original and the translated version of the texts. Supplementary Fig. S3 shows word clouds of the most common terms in the translated documents.

### Annotation of primary tumor sections

For training AI models on WSIs using supervised learning, the annotation or segmentation of present tumor regions is usually required [22]. WSIs often contain large areas of tissue that might be irrelevant or even misleading for the corresponding task. We sparsely annotated representative tumor areas in the primary tumor sections using QuPath. To this end, we manually selected one or several regions of interest representing the tumor’s histology while avoiding areas that contain artifacts, white background, or healthy tissue such as muscular or glandular tissue. This approach is based on the protocol for the analysis of deep texture representations [41]. An exhaustive annotation of all present tumor regions or distinct tissue types was not possible due to time constraints. We provide the resulting polygon annotations in ”.geojson” format to enable effortless extraction of tumor tiles for future works.

### Multimodal patient vectors

We created multimodal patient vectors for two purposes. First, the vectors were used to determine a dataset split for training and testing. Second, they were used to train models to predict outcomes or treatment choices. To this end, we created embeddings that condensed data from each modality and concatenated them to a single vector per patient.

We encoded the clinical and pathological features using different techniques based on their type. Binary encoding was applied for features such as lymphatic, vascular, or perineural invasion, the patient’s sex, or the presence of carcinoma in situ. The pT stage and pN stage were considered ordinal features and transformed into consecutive labels. Categorical features such as primary site or histologic type were assigned labels and were later one-hot encoded. For integrating laboratory parameters, we used the raw values of the hematology group, i.e. the complete blood count.

The ICD codes, extracted from surgery reports, provide a more detailed classification of the disease than the available structured data does. The sequence of ICD codes for each patient was considered a sentence and converted to vectors using a bag-of-words model, inspired by the bag-of-disease-codes approach by Placido et al. [42]. To this end, the first four characters of each ICD code were used. Codes covered by less than three patients were discarded.

The structured pathological data did not contain any information about the immune response of each patient. To include this information, we performed a quantitative analysis of TMAs using the open-source software QuPath (version 0.4.3) [43]. The density of T lymphocytes has been shown to be a prognostic marker [44, 45]. Inspired by the Immunoscore [24, 46], we computed the density of CD3- and CD8-positive cells in the tumor center and invasion front per tumor area. To this end, we used QuPath to de-array the TMAs and match the tissue cores with patient IDs. Next, tissue detection was performed using thresholding. Strong artifacts were manually removed from the detected regions. Using QuPath’s positive cell detection feature, we obtained the positive cell count per mm^2^ tumor area. Supplementary Fig. S13A shows exemplary TMA cores with detected positive cells and Supplementary Fig. S13B the respective cell densities. The distribution of the densities is shown in Supplementary Fig. S13C.

The single-modality vectors for each patient were finally concatenated to a multimodal vector with a length of 103. We used UMAP to visualize the multimodal patient vectors in 2D. Beforehand, one-hot encoding was performed for categorical features, missing values were imputed, and z-score normalization was applied to ordinal and numeric features, i.e. the values were centered around the mean with unit variance. The axes were normalized to the range between zero and one.

### Dataset split using a genetic algorithm

We aimed to provide a training dataset and a test dataset that is suitable to test any AI algorithm for its generalizability. We aimed for our test dataset to fulfill the following criteria proposed by Wagner et al. [18]: First, the data should be split at a patient level. Second, both datasets should follow a similar distribution of target classes, in this case, the recurrence and survival status. We created two distinct dataset splits, each into 80% training and 20% test data. The first split should follow the distribution of the training dataset concerning relevant characteristics, by including information from different modalities. The second should be out of distribution and contain outlier cases. To create both splits, we used evolutionary optimization [47].

We implemented a genetic algorithm, where each individual represented a possible split by a vector of zeros (patients assigned to training) and ones (patients assigned to test). The objective of the genetic algorithm was to maximize the fitness of an individual, i.e. of a split with N test points. Before computing the fitness of each split, missing values were imputed and categorical features were subsequently one-hot encoded. A penalty was subtracted from the fitness to achieve a class-balanced split. This penalty was defined as the sum of differences between each class distribution *d* = 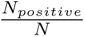 overall and in the current test dataset. Considering recurrence and survival status as target classes, the number of classes was *C* = 2 in our case. The penalty for C classes was weighted by a weight α. A similar approach was introduced by Florez-Revuelta who used a genetic algorithm to split multi-label data while maximizing the similarity between class distributions [48]. We calculated the fitness of an individual as follows:

For the in-distribution split, the fitness of an individual was defined as the sum of cosine distances from each test point x*_i_* to its nearest neighboring test point x*_i,nn_*:

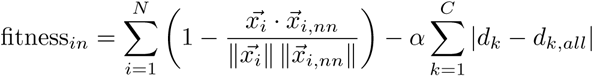

For the out-of-distribution split, we calculated the sum of cosine distances between all pairs of test points *x*:

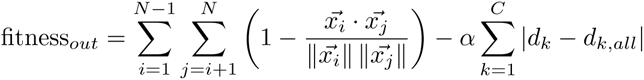

The population size was set to 10,000 and the genetic algorithm was terminated after 50 iterations with no further improvement. The population was iteratively updated using parent selection (tournament selection with elitism) and one-point crossover with inversion mutation until convergence. The genetic algorithm was only applied to patients with complete patient vectors. However, for some patients not all required modalities were available. These were subsequently assigned to the training dataset. The final splits were summarized as a list of patient IDs in JSON format.

### Outcome prediction for distinct dataset splits

For training Machine Learning models to predict recurrence or survival, three different data splits were used. The first split defined ”in distribution” cases as test data, the second split defined ”out of distribution” data as test data, and the third split defined cases with oropharyngeal cancer as test data (see Figure 2D. For survival prediction (see Figure 2E), cases with non-tumor-specific death were excluded. All other cases, including those with unknown causes of death, were considered. The class labels correspond to the survival status, i.e. ”living” and ”deceased”. Binary class labels were also defined for recurrence prediction (see Figure 2E). The classes were defined as (i) patients who had no recurrence and survived at least three years and (ii) patients who had a recurrence within three years.

As recommended by Huang et al., we applied an early fusion approach as an initial strategy, i.e. we created the multimodal patient vectors and trained a single model [19]. We used three different train-test splits of the dataset, namely the in-distribution and out-of-distribution datasets created using the genetic algorithm. Another split was created by assigning all laryngeal carcinomas to the test dataset. We used the Synthetic Majority Oversampling Technique (SMOTE) to handle class imbalance [49]. One classifier was trained and tested for each of the three splits (see Figure 2D).

### Treatment prediction using Machine Learning

To explore the ability of an AI model to suggest whether adjuvant therapy is needed or not, we split the dataset in the following way. Patients who had no adjuvant therapy but were deceased or had a recurrence, metastasis, or progress were assigned to the hold-out test dataset. All remaining cases were assigned to the training dataset, as shown in Figure 3A. We chose this setting to explore if an AI model could potentially identify patients who did not receive but would have needed adjuvant therapy. In identifying deceased patients, we considered the overall survival as the cause of death was not available for all cases. The class labels were defined as ”no adjuvant therapy used” and ”adjuvant therapy used”.

We used the single-modality vectors (clinical, pathological, blood, ICD codes, TMA cell densities) individually and their combination (multimodal patient vectors) to train Random Forest classifiers. For both single-modal and multimodal data, we used 10-fold cross-validation and reported the average ROC curve along with the AUC score. To handle the class imbalance problem, we applied SMOTE [49]. To avoid data leakage, we ensured that missing value imputation and normalization were performed in each iteration using statistics of the current training folds. To investigate the most important features, we computed SHAP values and visualized them for the ten most relevant features in a summary plot [23]. Finally, a classifier was trained on the full training dataset of multimodal patient vectors and the predictions for the test dataset were obtained. The training was performed for five iterations and the resulting ROC curves and AUC scores were averaged.

### Treatment prediction using Deep Neural Networks

Aiming to integrate histologic features into a model for treatment prediction, we used the same dataset split as before (see 3A) and trained a Convolutional Neural Network. To this end, we extracted features from TMAs stained using all eight available markers. Each slide image contains tissue cores of several patients. To map these cores to patient IDs, we de-arrayed the TMAs using QuPath and imported the TMA maps. Next, we extracted a single tile from the center of each TMA core. As for most patients, two cores and eight markers were available, resulting in 16 tiles per patient. Every tile was fed to a feature extractor to obtain an embedding vector of length 256. To this end, we used the feature extractor implemented by Komura et al. [27] which computes a gram-matrix of feature maps obtained from convolutional layers in the network and converts it to a one-dimensional embedding. We used a VGG16 as a feature extractor pre-trained on ImageNet and obtained features from the layer ”block3 conv3” [25, 26]. Next, we stacked the extracted image features and multimodal patient vectors to obtain a 2D embedding for each patient. Min-max scaling was applied to the image features using the minimum and maximum value computed from all image features in the training dataset. We trained a custom CNN on the image-like embeddings and performed a grid search to tune its hyperparameters. The approach of encoding and stacking multimodal features into a single source suitable for training CNNs was inspired by Nawaz et al. who fused image and text embeddings to improve classification performance [50].

We applied 10-fold cross-validation and reported ROC curves. A final model was trained on the full dataset and test predictions were obtained. To visually explain predictions, SHAP image plots were created for test samples [51]. As background samples for the SHAP algorithm, 100 random training samples were used.

### Data analysis

Overall survival curves were estimated using the Kaplan-Meier method [52]. The analysis considered the time between the initial diagnosis and death or the end of follow-up. Patients who were alive at the end of the follow-up were censored. We computed overall survival curves for all patients and for patients grouped by different characteristics, see Supplementary Fig. S6. For estimating the recurrence-free survival (see Fig. 3E), any occurrence of metastasis, progress, recurrence, or death was considered as an event and the duration was defined as the time between the first treatment (surgery) and the event.

The clinical data includes various events, such as treatments, progress of the disease, diagnosis of metastases, recurrence, and death or end of follow-up. We visualized the timelines of these events, see Supplementary Fig. S5.

### Statistics and Evaluation

The performance of classifiers was reported using ROC curves and corresponding AUC scores. To compute ROC curves and AUC scores, ML models were either trained and evaluated five times (see Figure 2E) or trained using 10-fold cross-validation (see Figures 3B and 4D). The ROC curves and AUC scores were then averaged over the iterations or the ten folds, respectively.

To evaluate the results of classifiers trained to predict adjuvant treatment, Kaplan-Meier curves were estimated and compared. To this end, we split the test cases into two groups based on the predicted classes. The first group contained cases where no adjuvant treatment was predicted (probability for adjuvant therapy recommendation below or equal to 0.5) and the second group contained cases where adjuvant treatment was predicted (probability above 0.5). Kaplan-Meier curves were estimated separately for the two groups. We used a log-rank test to compare survival curves and reported whether the p-value was below the significance level of 0.05 (*), 0.001 (**), or 0.0001 (***).

We applied the Wilcoxon-Mann-Whitney test to compare the distribution of CD3-positive and CD8-positive cell density of patients grouped by recurrence and survival status, as shown in Supplementary Fig. S8.

## Data Availability

The dataset is publicly available at https://hancock.research.fau.eu/.

## Data availability

The HANCOCK dataset is publicly available at https://hancock.research.fau.eu/. An overview of the dataset, including the number and format of files, is shown in Supplementary Fig. S14.

## Code availability

Code for data exploration, processing histologic images, feature extraction, generating data splits, outcome prediction, and adjuvant treatment prediction is available at https://github.com/ankilab/HANCOCK MultimodalDataset.

## Supplementary information

The supplement contains the Supplementary Figures S1-S14 and the Supplementary Tables S1-S4.

## Acknowledgments

This work was funded in part by the Federal Ministry of Education and Research (BMBF) to AOG and ME (01KD2211B) and to AMK (01KD2211A). We thank Mohammadhamed Mirbagheri for his excellent technical assistance.

