## Supplemental Material for "A multimodal dataset for precision oncology in head and neck cancer"

#### Supplementary Figures

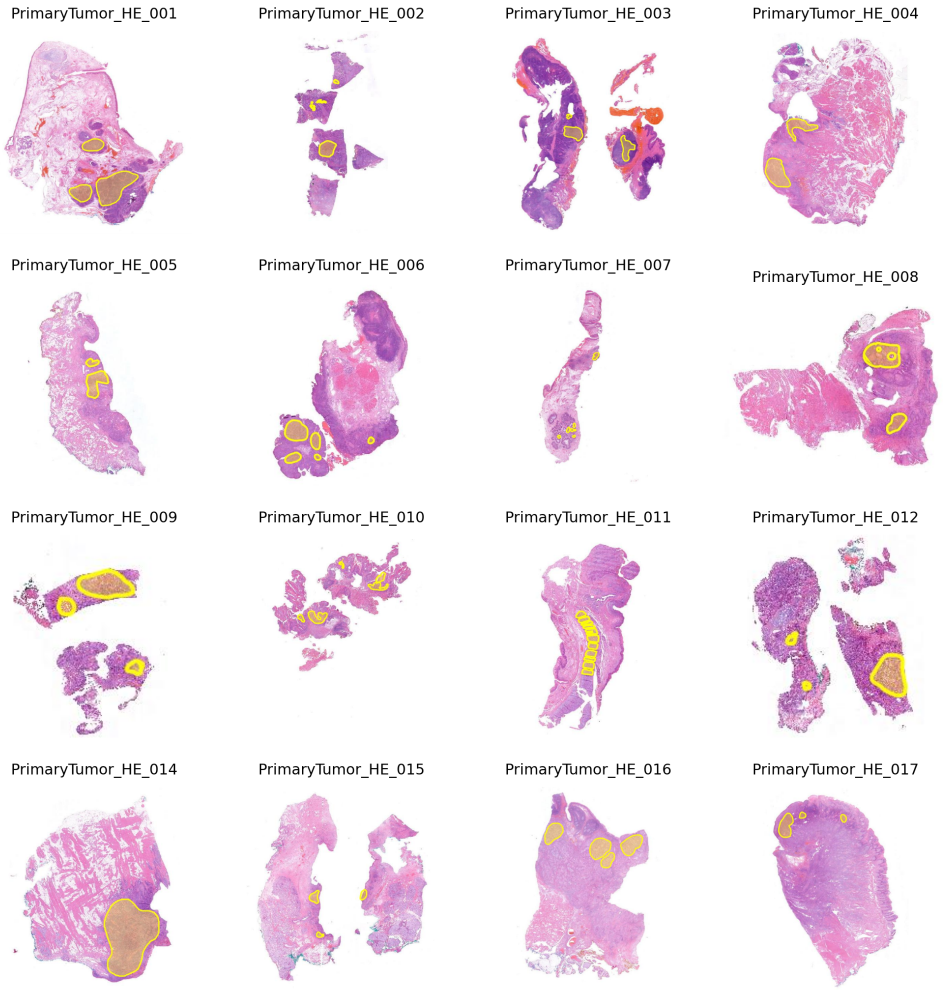


**Figure S1** Whole Slide Images of the primary tumor from 16 patients. Manual annotations of representative tumor regions are highlighted in yellow.

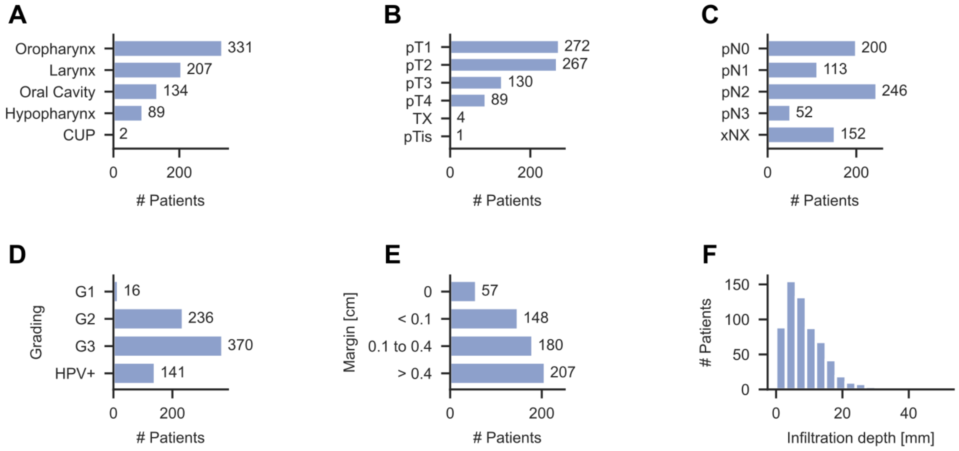


**Figure S2** Summary of pathologic data. (A) Number of cases per primary tumor site. There were two cases with cancer of unknown primary (CUP). (B) Cases per pathological T stage, where more specific stages such as pT1a, pT1b were grouped to pT1. (C) Cases per pathological N stage, where more specific stages such as pN2a, pN2b were grouped to pN2. (D) Number of cases vs. grading, where HPV-associated carcinomas were not graded. (E) Distance to closest resection margin, divided into four groups. (F) Histogram showing the distribution of tumor infiltration depth.

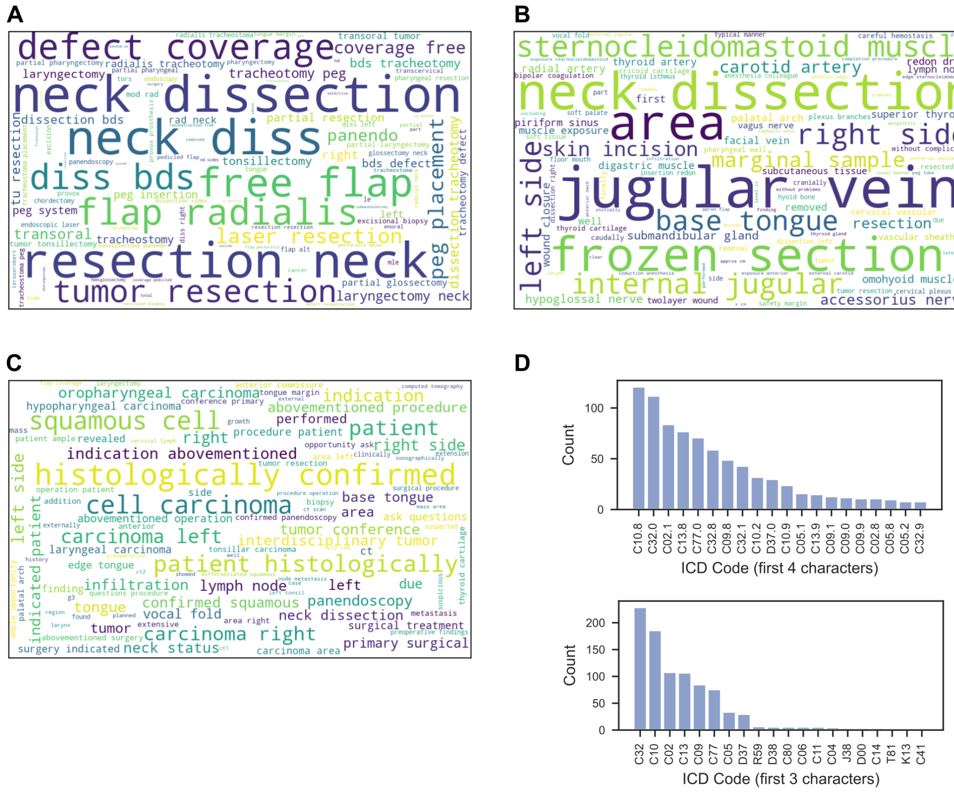


**Figure S3** Visualization of text data translated to English and frequency of ICD codes (International Statistical Classification of Diseases and Related Health Problems) of the German version ICD-10-GM. (A) Wordcloud of most frequent terms in short descriptions of surgeries. (B) Wordcloud of the most frequent terms in the surgery reports. (C) Wordcloud of the most frequent terms in medical histories. (D) Frequency of the top 20 ICD codes, considering the first 4 or 3 characters.

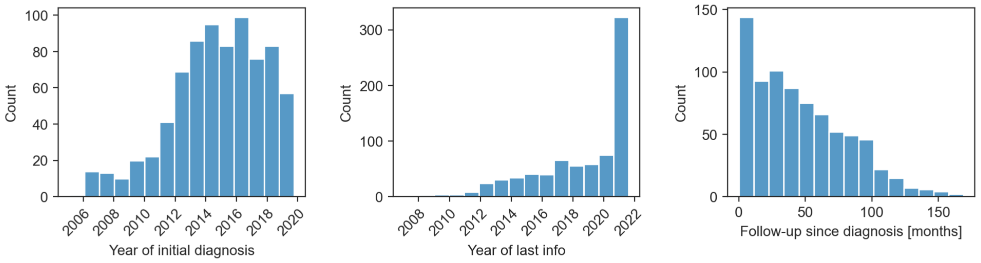


**Figure S4** Histograms showing how many patients were diagnosed between 2006 and 2020 along with the years in which the follow-ups ended, and the follow-up time from diagnosis to last information.

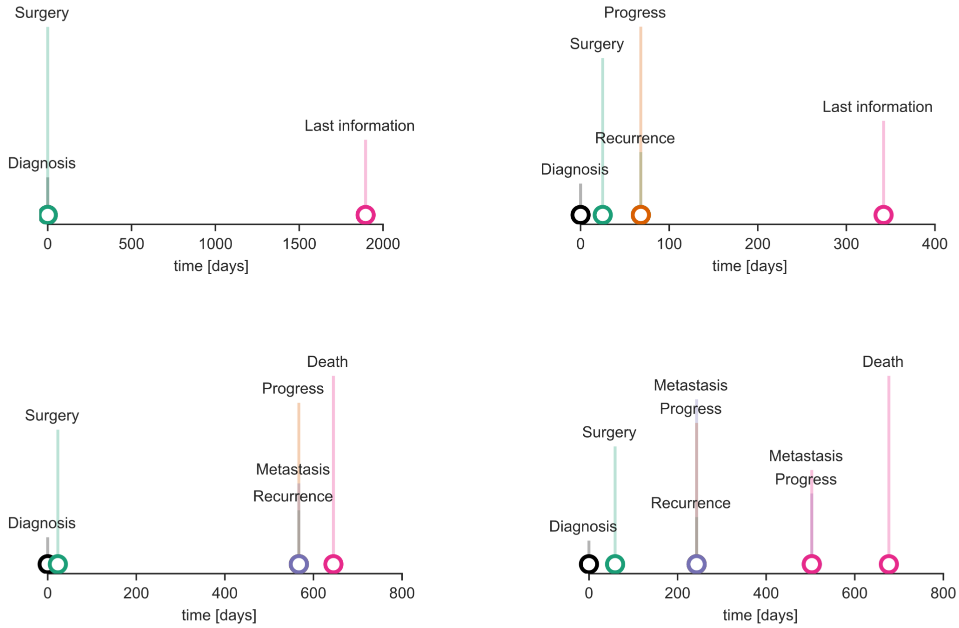


**Figure S5** Event timelines of four patients. In the top left example, the diagnosis was histologically confirmed externally, so the unknown diagnosis date was set to the surgery date. Considered events were diagnosis, surgery, progress, recurrence, metastasis, and death. Timestamps of possible adjuvant treatments were not included in the dataset as they were not available for all patients.

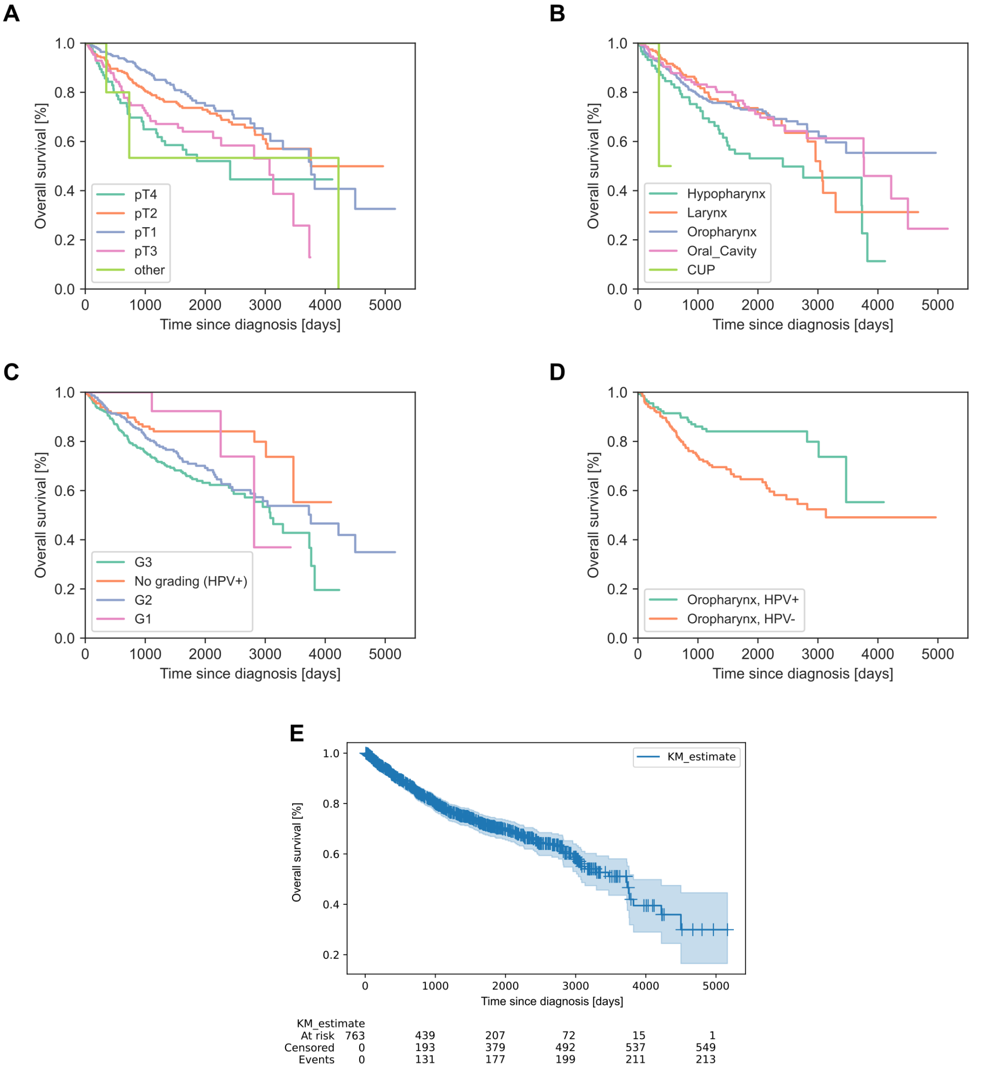


**Figure S6** Kaplan-Meier curves of overall survival. (A) Overall survival of patients grouped by pathological T stage. (B) Overall survival of patients grouped by grading. HPV-associated carcinomas were assigned to a separate group as these were not graded. (C) Overall survival of patients grouped by primary tumor site. The two cases with cancer of unknown primary are denoted as CUP. (D) Overall survival of patients with oropharyngeal carcinoma, grouped by HPV status (p16). (E) Overall survival with censored patients (+), the confidence interval, and the number of patients censored and at risk at different time points.

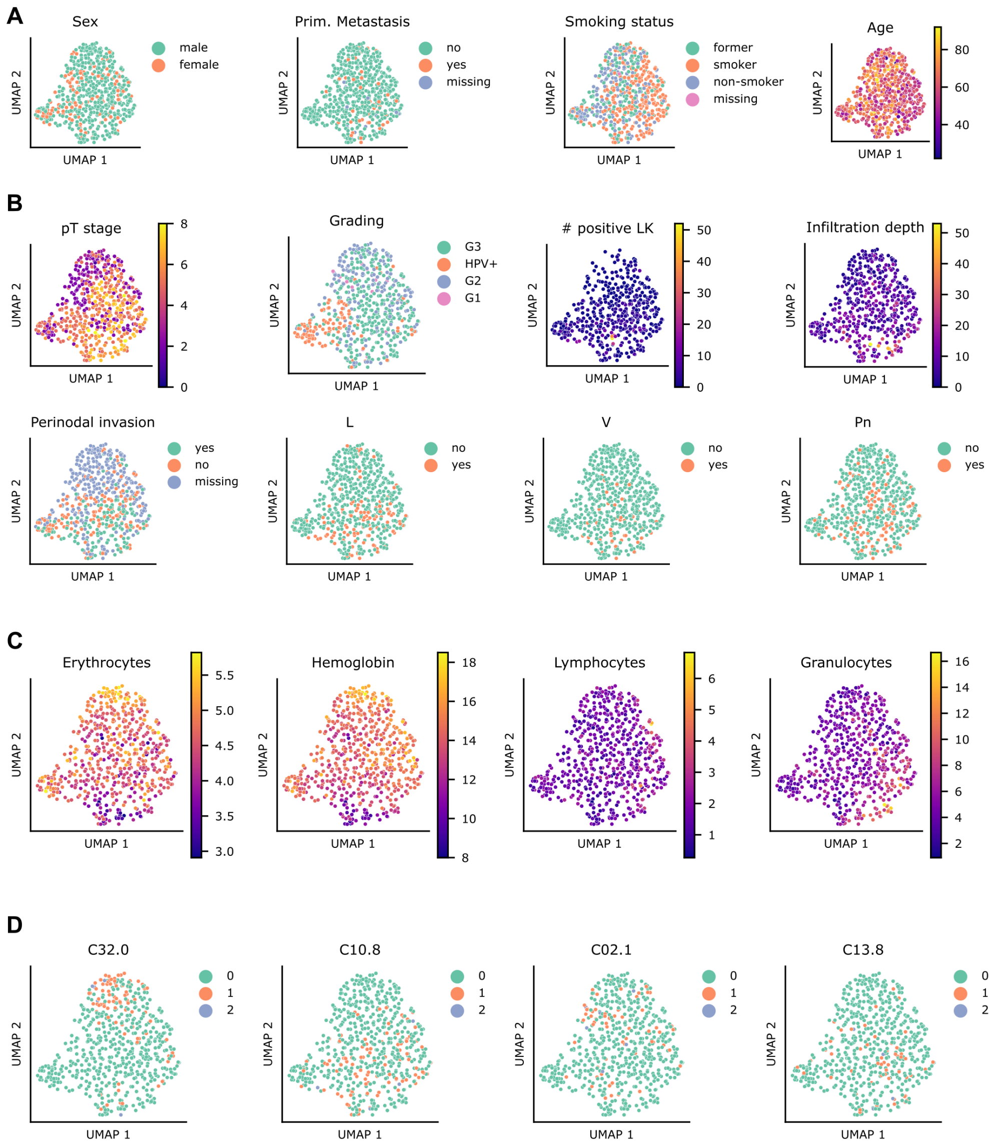


**Figure S7** Two-dimensional representation of the multimodal patient vectors, colored by different patient characteristics. (A) Demographics. (B) Pathology. (C) Blood parameters. (D) ICD Codes.

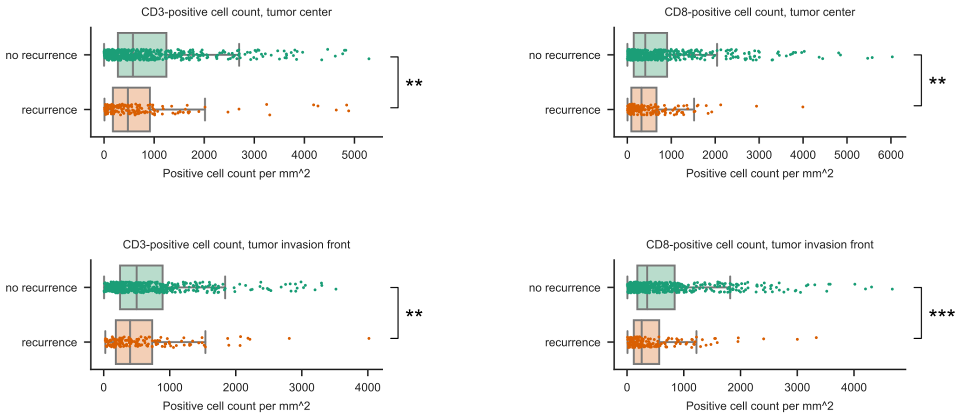


**Figure S8** The distribution of CD3- and CD8-positive cell counts per $mm^2$ were significantly greater for patients with no recurrence than for patients with recurrence. This applied for both the tumor center and tumor invasion front. We used the Wilcoxon-Mann-Whitney test. Two asterisks (**) denote a p-value below 0.01 and three (***) denote a p-value below 0.001.

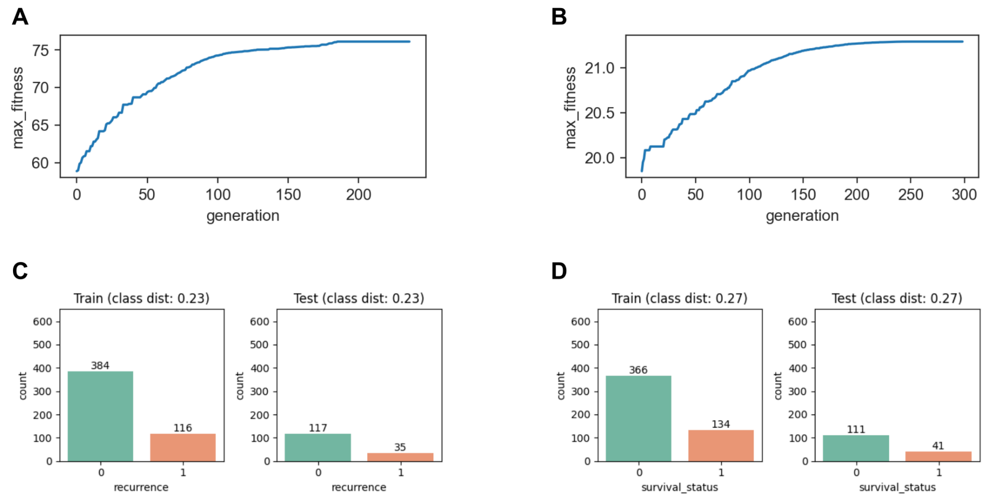


**Figure S9** Fitness and class distributions of the genetic algorithm. (A) Maximum fitness for each generation of the genetic algorithm for the in-distribution dataset split. (B) Maximum fitness for each generation of the genetic algorithm for the out-of-distribution dataset split. (C) Distribution of the target class ”recurrence” of the training and test dataset at the last generation. (D) Distribution of the target class ”survival status” at the last generation.

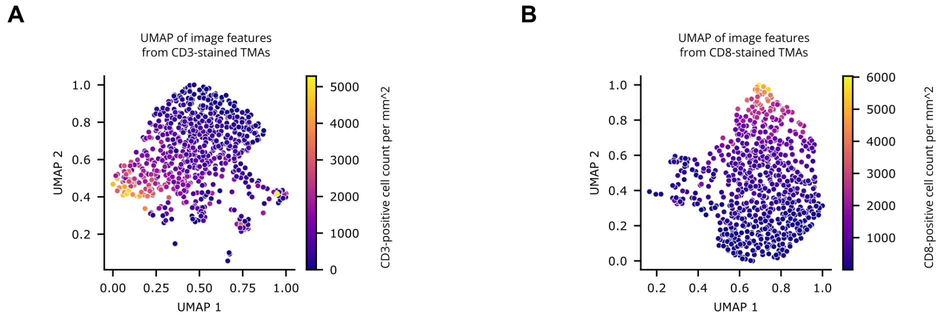


**Figure S10** Visualization of image features and cell densities using UMAP. (A) Two-dimensional representation of features extracted from CD3-stained TMAs, colored by calculated CD3-positive cell density. (B) Two-dimensional representation of features extracted from CD8-stained TMAs, colored by calculated CD8-positive cell density.

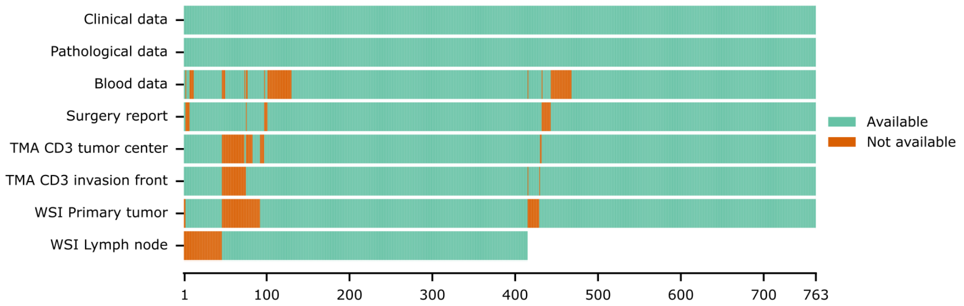


**Figure S11** Visualization of the available data. The red areas show for how many patients the individual data types are missing. For TMAs, The availability of TMA cores stained with CD3 are shown as an example. For most patients who are missing a primary tumor slide, a lymph node slide is available instead. Lymph node WSIs are only available for patients with lymph node metastases.

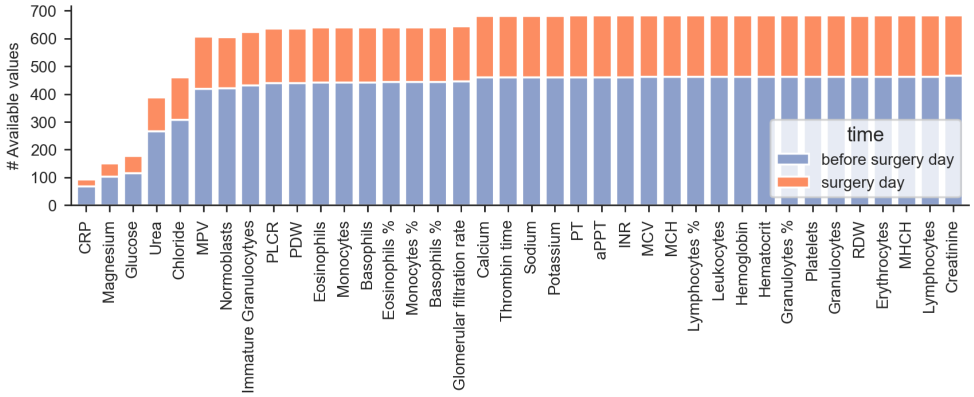


**Figure S12** Number of available measurements per parameter, after selecting a single measurement for each parameter and patient. Available pre-operative measurements are highlighted in blue. If this was not available, a measurement from the surgery day (highlighted in red) was selected instead.

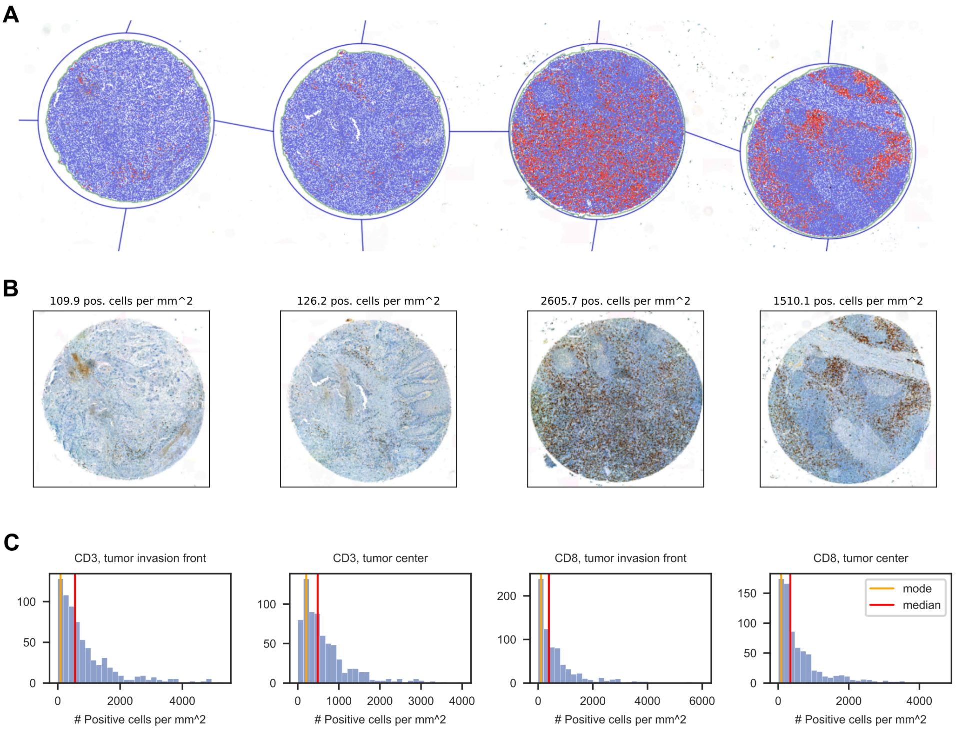


**Figure S13** Analysis of cells in Tissue Microarray (TMA) cores. (A) Four exemplary TMA cores with CD8 marker from the tumor invasion front. QuPath was used to de-array the TMA with the core diameter set to 1,9mm. Subsequently, positive cell detection was performed to detect CD8-positive cells. Positive cells are highlighted red and negative cells blue. (B) Number of CD8-positive cells per $mm^2$ for each core. (C) Histograms showing the distribution of CD3- and CD8-positive cell counts for all patients with available data. The mode and median values are highlighted.

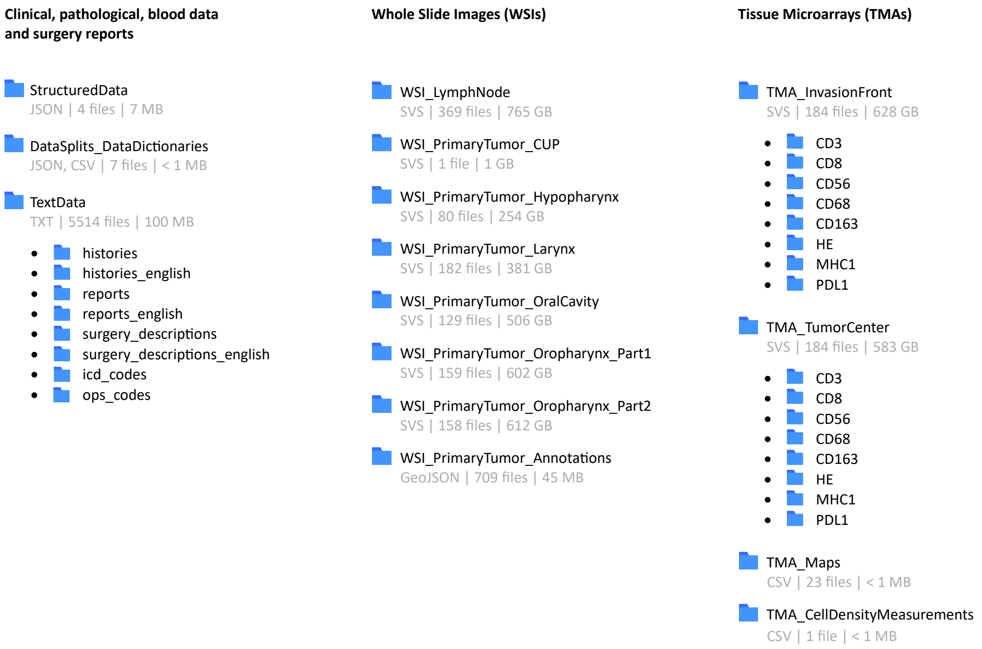


**Figure S14** Overview of the dataset, including file formats, numbers of files, and file sizes. The file formats in the dataset are JavaScript Object Notation (JSON), Comma-separated values (CSV), Plain text (TXT), Aperio SVS, and GeoJson. The total size of the dataset is 4.23 terabytes.

### Supplementary Tables

| Field | Description | Values | Data type |
|---|---|---|---|
| patient_id | Patient identifier, randomly assigned | ”001” to ”763” | string |
| year_of_initial_diagnosis | Year in which the patient was initially diagnosed with cancer | | integer |
| age_at_initial_diagnosis | Patient age in years | | integer |
| sex | Patient sex | female, male | string |
| smoking_status | First known smoking status of the patient | non-smoker, former, smoker | string |
| primarily_metastasis | Presence of metastases at the time of diagnosis | no, yes | string |
| survival_status | Survival status at the time of the last known information | living, deceased | string |
| survival_status_with_cause | Survival status with cause of death, if documented | living, deceased, deceased not tumor specific, deceased tumor specific | string |
| days_to_last_information | Time from diagnosis to last information or death, in days | | integer |
| first_treatment_intent | Intent of the first treatment, only patients with curative treatment intent were included | curative | string |
| first_treatment_modality | Modality of the first treatment, only patients with local surgery were included local surgery | | string |
| first_treatment_descr_german | Short description of the first treatment in German | | string |
| first_treatment_descr_english | Short description of the first treatment, translated to English using ChatGPT-3.5 | | string |
| days_to_first_treatment | Time from diagnosis to first treatment, in days, is equal to zero if the diagnosis was confirmed externally | | integer |
| adjuvant_treatment_intent | Intent of adjuvant treatment, if applied | curative, palliative | string |
| adjuvant_radiotherapy | Use of systemic radiotherapy as adjuvant treatment | no, yes | string |
| adjuvant_rad_modality | Modality of adjuvant radiotherapy, if applied | brachytherapy, percutaneous radiotherapy | string |
| adjuvant_systemic_therapy | Use of systemic radiotherapy as adjuvant treatment | no, yes | string |
| adjuvant_syst_modality | Modality of systemic radiotherapy, if applied | cisplatin, docetaxel, carboplatin, fluorouracil, cetuximab, pembrolizumab | string |
| adjuvant_rct | Use of radiochemotherapy as adjuvant treatment | no, yes | string |
| recurrence | Occurrence of a locoregional recurrence of cancer | no, yes | string |
| days_to_recurrence | Time from diagnosis to recurrence, if any, in days | | integer |

| | | | |
|---|---|---|---|
| progress_1 | Progress of the cancer, for example distant metastasis, late recurrence, or tumor growth | no, yes | string |
| days_to_progress_1 | Time from diagnosis to first progress, if any, in days | | integer |
| progress_2 | Second progress of the cancer, for example distant metastasis, late recurrence, or tumor growth | no, yes | string |
| days_to_progress_2 | Time from diagnosis to second progress, if any, in days | | integer |
| metastasis_1_locations | Localization of first known distant metastases, if present | Adrenal, Bones, Brain, Liver, Lungs, Lymph Nodes, Other Organs, Peritoneum, Pleura, Skin, Soft Tissue, Spleen | string |
| days_to_metastasis_1 | Time from diagnosis to first metastases, if present, in days | | integer |
| metastasis_2_locations | Localization of distant metastases at a second time point, if present | Refer to metastasis_1_locations | string |
| days_to_metastasis_2 | Time from diagnosis to second metastases, if present, in days | | integer |
| metastasis_3_locations | Localization of distant metastases at a third time point, if present | Refer to metastasis_1_locations | string |
| days_to_metastasis_3 | Time from diagnosis to third metastases, if present, in days | | integer |
| metastasis_4_locations | Localization of distant metastases at a fourth time point, if present | Refer to metastasis_1_locations | string |
| days_to_metastasis_4 | Time from diagnosis to fourth metastases, if present, in days | | integer |

**Table S1**: Clinical data dictionary.

| Field | Description | Values | Data type |
|---|---|---|---|
| patient_id | Patient identifier, randomly assigned | ”001” to ”763” | string |
| primary_tumor_site | Site of the primary tumor or CUP (cancer of unknown primary) | Hypopharynx, Larynx, Oral_Cavity, Oropharynx, CUP | string |
| pT_stage | Pathological T stage, with unknown stage TX | pT1, pT1a, pT1b, pT2, pT3, pT4a, pT4b, pTis, TX | string |
| pN_stage | Pathological N stage, with unknown stage NX | pN0, pN1, pN1a, pN2, pN2a, pN2b, pN2c, pN3, pN3b, NX | string |
| grading | Tumor grading, HPV-positive oropharyngeal carcinoma was not graded | G1, G2, G3, HPV_OSCC | string |
| hpv_association_p16 | HPV (human papillomavirus) status as indicated by p16 positivity (=aberrant overexpression), only patients with HPV-positive oropharyngeal carcinoma were tested | negative, positive, not_tested | string |
| histologic_type | Histologic type of the primary tumor | Mucoepidermoid_Carcinoma, Neuroendocrine_Carcinoma, SCC_Acantholytic, SCC_Basaloid, SCC_Conventional-Keratinizing, SCC_Conventional-NonKeratinizing, SCC_Lymphoepithelial, SCC_Sarcomatoid | string |
| number_of_positive_lymph_nodes | Number of resected lymph nodes with present cancer cells | | integer |
| number_of_resected_lymph_nodes | Number of resected lymph nodes | | integer |
| perinodal_invasion | Perinodal invasion | no, yes | string |
| lymphovascular_invasion | Lymphovascular invasion (L) | no, yes | string |
| vascular_invasion | Vascular invasion (V) | no, yes | string |
| perineural_invasion_Pn | Perineural invasion (Pn) | no, yes | string |
| resection_status | Resection status after first treatment, with unknown status RX | R0, R1, R2, RX | string |
| resection_status_carcinoma_in_situ | Resection status for carcinoma in situ | CIS_absent, Ris0, Ris1 | string |
| carcinoma_in_situ | Presence of carcinoma in situ | no, yes | string |
| closest_resection_margin_in_cm | Distance to closest resection margin, in cm. Distances smaller than 0.1 cm are denoted with “<0.1” | | string |
| infiltration_depth_in_mm | Infiltration depth of the tumor, in mm | | float |

**Table S2**: Pathological data dictionary.

| Field | Description | Values | Data type |
|---|---|---|---|
| patient_id | Patient identifier, randomly assigned | ”001” to ”763” | string |
| value | Measured value of the analyte | | float |
| unit | Unit | % | string |
| | | fl, | |
| | | g/dl, | |
| | | mg/dl, | |
| | | mg/l, | |
| | | ml/min, | |
| | | mmol/l, | |
| | | pg, | |
| | | s, | |
| | | $\times 10^3/\mu$l | |
| | | $\times 10^6/\mu$l | |
| analyte_name | Short name of the analyte | Basophils, | string |
| | | Basophils %, | |
| | | CRP, | |
| | | Calcium, | |
| | | Chloride, | |
| | | Creatinine, | |
| | | Eosinophils, | |
| | | Eosinophils %, | |
| | | Erythrocytes, | |
| | | Glomerular filtration rate, | |
| | | Glucose, | |
| | | Granulocytes, | |
| | | Granulocytes %, | |
| | | Hematocrit, | |
| | | Hemoglobin, | |
| | | INR, | |
| | | Immature Granulocytyes, | |
| | | Leukocytes, | |
| | | Lymphocytes, | |
| | | Lymphocytes %, | |
| | | MCH, | |
| | | MCV, | |
| | | MHCH, | |
| | | MPV, | |
| | | Magnesium, | |

| | | | | |
|---|---|---|---|---|
| | | Monocytes, | | |
| | | Monocytes %, | | |
| | | Normoblasts, | | |
| | | PDW, | | |
| | | PLCR, | | |
| | | PT, | | |
| | | Platelets, | | |
| | | Potassium, | | |
| | | RDW, | | |
| | | Sodium, | | |
| | | Thrombin time, | | |
| | | Urea, | | |
| | | aPPT | | |
| LOINC_code | LOINC (Logical Observation Identifiers Names and Codes) code | 26444-0, | | string |
| | | 30180-4, | | |
| | | 1988-5, | | |
| | | 2000-8, | | |
| | | 2075-0, | | |
| | | 2160-0, | | |
| | | 26449-9, | | |
| | | 26450-7, | | |
| | | 26453-1, | | |
| | | 33914-3, | | |
| | | 2345-7, | | |
| | | 30394-1, | | |
| | | 19023-1, | | |
| | | 20570-8, | | |
| | | 718-7, | | |
| | | 34714-6, | | |
| | | 38518-7, | | |
| | | 26464-8, | | |
| | | 26474-7, | | |
| | | 26478-8, | | |
| | | 28539-5, | | |
| | | 30428-7, | | |
| | | 28540-3, | | |
| | | 28542-9, | | |
| | | 2601-3, | | |
| | | 26484-6, | | |
| | | 26485-3, | | |
| | | 33990-3, | | |
| | | 32207-3, | | |

| | | | |
|---|---|---|---|
| | | 48386-7, | |
| | | 5894-1, | |
| | | 26515-7, | |
| | | 2823-3, | |
| | | 30385-9, | |
| | | 2951-2, | |
| | | 3243-3, | |
| | | 3091-6, | |
| | | 3173-2 | |
| LOINC_name | LOINC Long Common Name | Basophils [#/volume] in Blood, | string |
| | | Basophils/100 leukocytes in Blood, | |
| | | C reactive protein [Mass/volume] in Serum or Plasma, | |
| | | Calcium [Moles/volume] in Serum or Plasma, | |
| | | Chloride [Moles/volume] in Serum or Plasma, | |
| | | Creatinine [Mass/volume] in Serum or Plasma, | |
| | | Eosinophils [#/volume] in Blood, | |
| | | Eosinophils/100 leukocytes in Blood, | |
| | | Erythrocytes [#/volume] in Blood, | |
| | | Glomerular filtration rate/1.73 sq M.predicted by Creatinine-based formula (MDRD), | |
| | | Glucose [Mass/volume] in Serum or Plasma, | |
| | | Granulocytes [#/volume] in Blood, | |
| | | Granulocytes/100 leukocytes in Blood by Automated count, | |
| | | Hematocrit [Volume Fraction] of Blood, | |
| | | Hemoglobin [Mass/volume] in Blood, | |
| | | INR in Blood by Coagulation assay, | |
| | | Granulocytes Immature/100 leukocytes in Blood, | |
| | | Leukocytes [#/volume] in Blood, | |
| | | Lymphocytes [#/volume] in Blood, | |
| | | Lymphocytes/100 leukocytes in Blood, | |
| | | Erythrocyte mean corpuscular hemoglobin [Entitic mass], | |
| | | Erythrocyte mean corpuscular volume [Entitic volume], | |
| | | Erythrocyte mean corpuscular hemoglobin concentration [Mass/volume], | |
| | | Platelet mean volume [Entitic volume] in Blood, | |
| | | Magnesium [Moles/volume] in Serum or Plasma, | |
| | | Monocytes [#/volume] in Blood, | |
| | | Monocytes/100 leukocytes in Blood, | |
| | | Normoblasts/100 leukocytes [Ratio] in Blood, | |
| | | Platelet distribution width [Entitic volume] in Blood by Automated count, | |
| | | Platelets Large/Platelets in Blood by Automated count, | |

| | | | |
|---|---|---|---|
| | | Prothrombin time (PT) actual/normal in Platelet poor plasma by Coagulation assay, | |
| | | Platelets [#/volume] in Blood, | |
| | | Potassium [Moles/volume] in Serum or Plasma, | |
| | | Erythrocyte distribution width [Ratio], | |
| | | Sodium [Moles/volume] in Serum or Plasma, | |
| | | Thrombin time in Platelet poor plasma by Coagulation assay, | |
| | | Urea [Mass/volume] in Serum or Plasma, | |
| | | Activated partial thromboplastin time (aPTT) in Blood by Coagulation assay | |
| group | Analyte group | Electrolytes - single valence, | string |
| | | Hematology, | |
| | | Mineral; bone; joint; connective tissue, | |
| | | OG, | |
| | | Protein, | |
| | | Renal function, | |
| | | Routine, | |
| | | Sugars/Sugar metabolism | |
| days_before_first_treatment | Number of days before the surgery, i.e. 0 corresponds to the surgery day and 1 corresponds to one day before the surgery | [0, 14] | int |

**Table S3**: Blood data dictionary.

| Personal identifier | Placeholder | # Replaced in surgery reports | # Replaced in medical histories |
|---|---|---|---|
| Name of a clinician e.g. surgeon | <CLINICIAN_NAME> | 941 | 14 |
| Name of the patient | <PATIENT_NAME> | 0 | 21 |
| Date or month | <[year]> | 4 | 136 |
| Name of a study | <STUDY_NAME> | 5 | 0 |
| Location | <LOCATION> | 3 | 3 |

**Table S4** Number of placeholders used in surgery reports and medical histories for de-identification. Patient names had already been masked in the reports but appeared in some medical histories. Dates in the text were replaced with the corresponding year, for example, <2024>.
